# Know Your Epidemic, Know Your Response: Covid-19 in the United States

**DOI:** 10.1101/2020.04.09.20049288

**Authors:** Alberto Ciancio, Fabrice Kämpfen, Iliana V. Kohler, Daniel Bennett, Wändi Bruine de Bruin, Jill Darling, Arie Kapteyn, Jürgen Maurer, Hans-Peter Kohler

**Author notes:** Corresponding authors: Alberto Ciancio and Hans-Peter Kohler.

## Abstract

We document that during the week of March 10–16, the Covid-19 pandemic fundamentally affected the perceptions of U.S. residents about the health risks and socioeconomic consequences entailed by the pandemic. During this week, it seems, “everything changed.” Not only did the pandemic progress rapidly across the United States, but U.S. residents started to realize that the threat was real: increasing Covid-19 caseloads heightened perceptions of infection risks and excess mortality risks, concerns about the economic implications increased substantially, and behavioral responses became widespread as the pandemic expanded rapidly in the U.S. In early to mid-March 2020, average perceptions about the coronavirus infection risks are broadly consistent with projections about the pandemic, while expectations about dying conditional on infection and expectations about Covid-19-related excess mortality during the next months are possibly too pessimistic. However, some aspects of Covid-19 perceptions are disconcerting from the perspective of implementing and sustaining an effective societal response to the pandemic. For instance, the education gradient in expected infection risks entails the possibility of having different perceptions of the reality of the pandemic between people with and without a college education, potentially resulting in two different levels of behavioral and policy-responses across individuals and regions. Unless addressed by effective health communication that reaches individuals across all social strata, some of the misperceptions about Covid-19 epidemic raise concerns about the ability of the United States to implement and sustain the widespread and harsh policies that are required to curtail the pandemic. Our analyses also reveal perceptions of becoming infected with the virus, and dying from Covid-19, were driven upwards by a rapidly increasing national caseload, and perceptions of the economic consequences and the adaptation of social distancing were affected by both national and state-level cases.

---

“Know your epidemic, know your response”, a rallying theme of HIV prevention with applicability to the Covid-19 pandemic, highlights the fact that individuals’ knowledge about infection risks are critical for their ability to respond to the pandemic with appropriate risk reduction strategies, and support the restrictions on individuals’ lives that are implemented to curtail the spread of the virus (1–4). Observers now recognize that the United States was late to adopt proactive measures to address COVID-19 in the early stages of the epidemic (5). The lack of early implementation of testing and social distancing may reflect in part that people did not perceive COVID-19 as a severe risk.

Using the Covid-19 focused questionnaire of the Understanding America Study(6) implemented during March 10–16, 2020, our study is the first to use weighted nationally-representative data to document individuals’ perceptions of the risks and consequences of the Covid-19 pandemic in the United States. The survey elicited from respondents probabilities of events—such as getting infected with coronavirus or dying in case of infection— on a scale from 0 to 100, and also asked respondents whether they had taken steps to stay away from others such as avoiding public spaces and canceling work or social activities (see Supplemental Materials). By March 16, the online survey had collected data from 5,414 persons.

Around mid-March, when the Covid-19 epidemic rapidly expanded in the United States, U.S. residents perceived on average a 20% chance of getting infected with the coronavirus (SARS-CoV-2) during the next three months (Fig. 1a and Table S1). These 3-months average infection risks of about one out of five persons are consistent with disease scenarios in which the containment of the disease is no longer possible and a large fraction of the U.S. population will ultimately get infected with the virus, while social distancing is able to flatten the curve and spread infections across an extended time period (7). Yet, underneath this 20% average is considerable heterogeneity: 21% of U.S. residents perceive zero risk of getting infected in the next 3 months, while 21.8% perceive a 0.1–10% risk, 17.6% a 10.1–20% risk, and 39.7% a 20% or higher chance. Only a small fraction, 2.4%, perceives a 3-month infection risk of 80% or higher (Table S2). Neither the average infection risk, nor the fraction perceiving fairly low or fairly high infection risks differ substantially between the three states that had already widespread community transmission during the week of March 10–16 (Washington, California, New York) and the rest of the United States (Table S3). There are also no significant gender differences (Fig. S1a). Important differences, however, emerge across educational categories and age groups, with more educated and younger persons perceiving higher infection risks (Fig. 1a and Fig. S1a). For example, U.S. residents in their 30s with a Bachelor’s degree or higher perceive a 50% higher chance of being infected than their lower educated peers. There are also differences by individuals’ sources of news: U.S. residents who use Fox News to learn about the coronavirus report a 5 percentage points lower probability of being infected than U.S. residents who use CNN (Table S3). These differences persist controlling for basic demographic characteristics as well as state and day fixed effects (Table S5).

**Figure 1:**
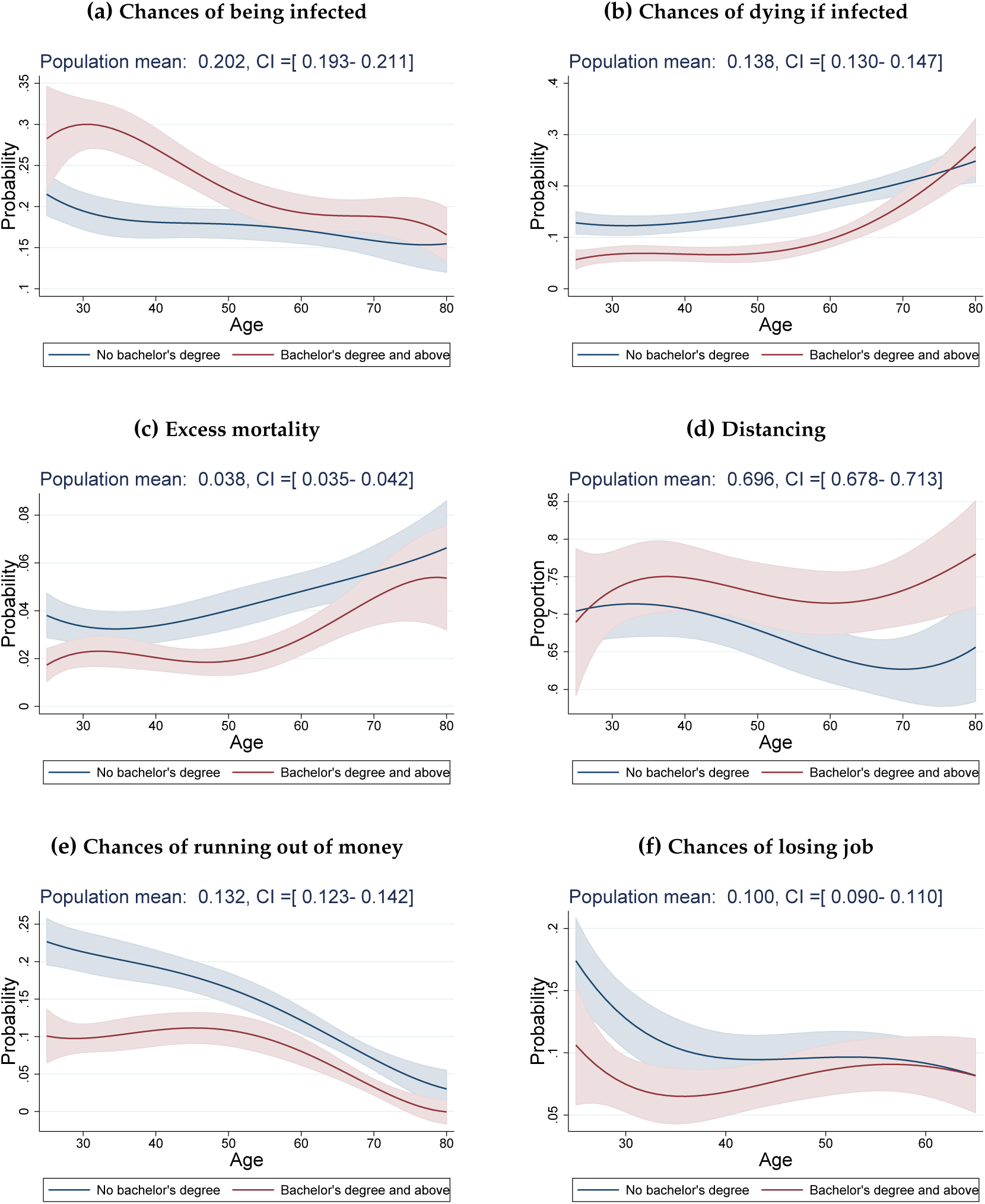
Differences by education level. *Notes:* Data coming from “Understanding America Study” (UAS) collected between March 10 and March 16, 2020. The plots show the marginal effects of an increase in age (1 year) on a) the chances of getting the virus within three months (top left), b) the chances of dying from the virus if infected (top right), c) excess mortality (middle left), d) whether individuals refrain from at least one social activity (middle right), e) the chances of running out of money because of the virus within three months (bottom left) and f) the chances of losing job within three months (bottom right). Effects for those with a Bachelor’s degree and above are represented by red lines and by blue ones for others. Marginal effects for individuals aged between 25 and 80 are the results of weighted regressions that include quartic polynomial in age. Shaded areas represent 95% confidence intervals (robust standard errors).

Individuals in the U.S. on average perceive a 14% chance of dying if infected with the coronavirus (Fig. 1b and Table S1), and this average perceived mortality rate exceeds ten-fold common estimates of the mortality risk due to Covid-19, while it is close to rates of death conditional on testing positive in areas where the health system is already failing and testing is restricted to the most ill patients (e.g., Lombardy in Italy). While the perceived mortality risk among U.S. residents from Covid-19 is relatively high, there is substantial variation across individuals: 23.6% perceive the chance of death if infected with coronavirus of zero while 39.6% perceive this risk as 0.1–10%, 13.2% as 10.1–20% and 23.6% as 20% or higher (Table S2). These different perceptions of mortality if infected are importantly related to education and age. For example, individuals with a Bachelor’s degree or higher estimate their chances of dying to be about 8 percentage points lower across primary adult ages than those with less education (Fig. 1b). This gradient in education diminishes among older persons (Fig. 1b), while they (accurately) perceive substantially greater mortality risks (Fig. S1b).

Combining the risk of infection with the risk of death conditional on infection provides individuals’ perceived excess mortality as a result of Covid-19. On average, individuals in the U.S. perceive a 3.8% chance of dying as a result of Covid-19 in the next three months (Fig. 1c and Table S1), representing a significant perception of excess short-term mortality. These perceptions vary widely, with 31.6% expecting zero excess mortality from Covid-19, and 12% expecting an excess mortality in the next three months of 10% or more (Table S2). Perceptions of the excess mortality from Covid-19 are lower for more educated persons and higher for older individuals (Fig. 1c), while there are no strong differences by gender (Fig. S1c).

The actual excess mortality in the U.S. as a result of Covid-19 remains very uncertain at this point (8–10) and will depend strongly on the success of social distancing and other disease containment efforts (7, 11). Nevertheless, our results suggest that individuals in the U.S. are probably too pessimistic in that regard: while people in the U.S. have average expectations about getting infected within three months that are broadly consistent with the projected infected population in intermediate scenarios of how the pandemic will unfold, on average they overestimate the mortality conditional on getting infected.

There are important temporal aspects in the perceptions of Covid-19 during the week of March 10–16, 2020 when the identified cases increased from 959 to 4,632, and the reported Covid-19 deaths increased from 28 to 85 (12). Importantly, our analyses of Covid-19 perceptions during March 10–16 show that the rapid surge in caseload at the country level increased the perceived chance of getting infected with coronavirus: a one-unit increase in the log number of Covid-19 cases in the U.S. leads on average to an increase of about 0.055 in the perceived chances of getting infected by the virus (Table 1, top panel), which corresponds to a 3.8 percentage points increase for each doubling of the national U.S. Covid-19 caseload (0.055 × ln(2)). Neither the increase in national Covid-19 cases, nor the increase in deaths, was positively associated with expectations about dying conditional on being infected with Covid-19 or the perceived excess mortality as a result of Covid-19 (Table 1 and Table S6). If anything, the increase in national cases had a negative (but imprecisely estimated) impact on the perceived probability of dying conditional on infection. This change in perceptions is consistent with a scenario where expanded testing across the country increasingly identifies milder cases with lower mortality risks conditional on infection.

**Table 1:** Effects of Covid cumulative cases on Covid perceptions and social distancing

|  | (1)<br>Get Covid | (2)<br>Die from Covid | (3)<br>Excess Mortality | (4)<br>Lose Job | (5)<br>Out of Money | (6)<br>Distancing |
| --- | --- | --- | --- | --- | --- | --- |
| Regressions with state fixed effects |  |  |  |  |  |  |
| US cases (log) | 0.055***<br>[0.029,0.081] | -0.017*<br>[-0.036,0.002] | 0.001<br>[-0.007,0.009] | 0.054***<br>[0.027,0.082] | 0.060***<br>[0.031,0.089] | 0.206***<br>[0.160,0.252] |
| Regressions with state and day fixed effects |  |  |  |  |  |  |
| State cases (log) | 0.015<br>[-0.017,0.047] | -0.001<br>[-0.021,0.018] | 0.001<br>[-0.011,0.013] | 0.021<br>[-0.010,0.051] | 0.034**<br>[0.001,0.067] | 0.054***<br>[0.014,0.094] |
| Observations | 5272 | 5270 | 5269 | 3245 | 5301 | 5287 |

*Notes:* The table shows the effects of the log of the number of US and state of residence Covid-19 cases on 1) the chances of getting the virus within three months, 2) the chances of dying from the virus if infected, 3) excess mortality, 4) the chances of losing job within three months, 5) the chances of running out of money because of the virus within three months and 6) whether individuals refrain from at least one social activity. The top panel shows regressions that include state fixed effects while the bottom panel shows regression that include both state and day of interview fixed effects. State cases are transformed using the inverse hyperbolic sine $\log(x + \sqrt{1 + x^2})$ which is similar to log but allows for zeros. Additional controls include age, gender and four categories of education. We use sample weights to make the survey representative of the U.S. population aged 18 and older. 95% confidence intervals in squared brackets are calculated using standard errors clustered at the state level. \* $p < 0.1$ , \*\* $p < 0.05$ , \*\*\* $p < 0.01$ . Data on perceptions and social distancing come from “Understanding America Study” (UAS) collected between March 10 and March 16, 2020. Data on the number of Covid-19 cases come from the Johns Hopkins University Center for Systems Science and Engineering.

As of March 10–16, U.S. residents had increasingly adopted social distancing measures to delay the spread of the virus: about 70% of people in the U.S. report to have taken steps to stay away from other persons (“social distancing”) (Fig. 1d and Table S1), and despite the widely-circulated images of young persons agglomerating on beaches and in bars, overall younger U.S. residents were at least as likely (based on self-reports) to have taken social distancing measures as older persons. Women are more likely to engage in social distancing (Fig. S1d), while educational differences only emerge among older people where college educated individuals were significantly more likely to practice social distancing than non-college educated individuals (Fig. 1d). Using CNN as a source of information on coronavirus is associated with a 10 percentage point higher chance of engaging in social distancing than using Fox News (Table S3).

Individuals in the U.S. perceive an average 10% chance to lose their job within three months, and a 13% chance to run out of money in the next three months as a result of Covid-19 (Fig. 1e,f and Table S1). These perceptions highlight staggering pessimistic expectations about the short-term negative economic consequences as a result of the pandemic. There are again clear differences in these perceptions across education levels, with individuals holding at least a Bachelor’s degree having about twice lower chances to run out of money at younger ages as a consequence of the virus outbreak as compared to others (Fig. 1e and Fig. 1f). Women are more concerned about these economic consequences than men (Fig. S1e and Fig. S1f).

Importantly, U.S. residents’ perceptions of the socioeconomic severity of the pandemic changed fundamentally during the week of March 10–16. During these seven days, the increase in cases at the national level induced people in the U.S. to revise upward the perceived likelihood of losing their job or running out of money as a result of Covid-19, and taking steps to stay away from others (Table 1). Our analyses show that each doubling of the U.S. caseload implied an increase of 4.2 percentage points increase in the chance of running out of money within three months, and a 14.3 percentage point increase in the likelihood that the respondents have taken steps to stay away from others. U.S. residents thus increasingly realized within merely seven days that Covid-19 might entail drastic socioeconomic consequences, such as losing one’s job or running out of money within three months. These perceptions of the socioeconomic consequences of the epidemic, and the adaptation of social distancing measures, were particularly pronounced in states with the most severe epidemic (Table 1, bottom panel). Specifically, the increase in caseload in a respondent’s state of residence accelerated worries about running out of money (2.4 percentage points for each doubling of cases) and increased social distancing (3.7 percentage points for each doubling of cases) above and beyond the increase in cases at the national level. The results persist in analyses that control for whether local authorities have implemented measures requiring social distancing.

## Ethical/Institutional Review Board Approval

The Understanding America Study (UAS) was approved by the IRB at the University of Southern California through Protocol USC UPIRB UP-14-00148. Analyses at the University of Pennsylvania were based on publicly available de-identified UAS data obtained from the UAS project webpage at https://uasdata.usc.edu/index.php. The analyses at the University of Pennsylvania were exempt from IRB research (Exemption #4). No human subject research was conducted as part of this paper at the University of Lausanne.

## Data Availability

Data available: data are publicly available at https://uasdata.usc.edu; code for duplicating the analyses based on the publicly-available UAS data, including the additional data on Covid-19 caseloads, will be made available as part of final the publication.

## Supplementary Material

The data used in this study come from the Understanding America Study (UAS) fielded between March 10 and 16, 2020. The UAS is a panel of households at the University of Southern California of approximately 8,500 respondents representing the entire United States population. Participants are invited to respond to the survey online. More information can be found here: https://uasdata.usc.edu/index.php. This paper uses only observations from March 10 to 16 and thus includes only 5,414 respondents. Each respondent answered the Covid module only once. Sample weights are survey-specific and are meant to make each survey data set representative of the U.S. population aged 18 and older.

- The outcome variables used in the analysis are derived from the following questions:
- On a scale of 0 to 100 percent, what is the chance that you will get the coronavirus in the next three months? If you’re not sure, please give your best guess.
- If you do get the coronavirus, what is the percent chance you will die from it? If you’re not sure, please give your best guess.
- What is the percent chance that you will lose your job because of the coronavirus within the next three months?
- What is the percent chance you will run out of money because of the coronavirus in the next three months?
- Which of the following have you done in the last seven days to keep yourself safe from coronavirus in addition to what you normally do?
- Which of the following information sources have you used to learn about the coronavirus in the past 7 days?

Respondents had to answer questions 1 to 4 using a scale from 0 to 100. In the analysis, we rescaled answers from these questions by dividing them by 100. Question 3 was asked only to respondents who had a job at the time of the interview. Regarding our social distancing measure (question 5), we consider that a respondent was refraining from at least one social activity if the person admitted to have done at least one of the following: “Canceled or postponed travel for work”, “Canceled or postponed travel for pleasure”, “Canceled or postponed work or school activities”, “Canceled or postponed personal or social activities”, “canceled a doctor’s appointment”, “avoided contact with people who could be high-risk”, “avoided public spaces, gatherings, or crowds”, “avoided eating at restaurants” or “worked or studied at home”.

**Table S1:** Summary Statistics

|  | All | 95% conf. interval |  | Women | 95% conf. interval |  | Men | 95% conf. interval |  | <60 | 95% conf. interval |  | >=60 | 95% conf. interval |  |
| --- | --- | --- | --- | --- | --- | --- | --- | --- | --- | --- | --- | --- | --- | --- | --- |
| 1) Get Covid | **.202** | .193 | .211 | **.196** | .185 | .208 | **.207** | .194 | .221 | **.216** | .205 | .228 | **.168** | .155 | .182 |
| 2) Die from Covid | **.138** | .130 | .147 | **.149** | .138 | .161 | **.127** | .115 | .138 | **.114** | .105 | .123 | **.195** | .178 | .211 |
| 3) Excess Mortality | **.038** | .035 | .042 | **.041** | .036 | .046 | **.035** | .030 | .040 | **.032** | .028 | .036 | **.052** | .045 | .060 |
| 4) Lose Job | **.100** | .090 | .110 | **.114** | .100 | .129 | **.085** | .073 | .098 | **.103** | .092 | .114 | **.083** | .064 | .101 |
| 5) Out of Money | **.132** | .123 | .142 | **.162** | .148 | .175 | **.101** | .089 | .113 | **.163** | .151 | .176 | **.061** | .052 | .071 |
| 6) Distancing | **.696** | .678 | .713 | **.724** | .702 | .747 | **.665** | .638 | .692 | **.709** | .688 | .731 | **.664** | .634 | .694 |

Notes: The table shows the weighted mean and corresponding 95% confidence interval for all U.S. residents, and by gender and age categories (below or above 60) of 1) the chances of getting the virus within three months, 2) the chances of dying from the virus if infected, 3) excess mortality, 4) the chances of losing job within three months, 5) the chances of running out of money because of the virus within three months and 6) whether individuals refrain from at least one social activity. We use sample weights to make the survey representative of the U.S. population aged 18 and older. Data on perceptions and social distancing come from “Understanding America Study” (UAS) collected between March 10 and March 16, 2020.

**Table S2:** Summary statistics of the continuous measures used in the analysis

|  | Get Covid | Die from Covid | Excess Mortality | Lose Job | Out of Money |
| --- | --- | --- | --- | --- | --- |
| Population mean | .202 | .138 | .038 | .100 | .132 |
| Population median | .100 | .038 | .003 | 0 | 0 |
| *Weighted frequencies* |  |  |  |  |  |
| Probability of 0 | .210 | .236 | .316 | .526 | .513 |
| Probability > 0 and $\leq 10$ | .218 | .396 | .567 | .199 | .172 |
| Probability > 10 and $\leq 20$ | .176 | .132 | .051 | .095 | .082 |
| Probability > 20 | .397 | .236 | .067 | .179 | .233 |
| Probability > 80 | .024 | .029 | .001 | .018 | .038 |

*Note:* The table shows the weighted mean and median of the five continuous measures we use in the analysis (top panel). The bottom panel presents some characteristics of the distribution of these measures by means of weighted frequencies. The data come from "Understanding America Study" (UAS) collected between March 10 and March 16, 2020.

**Table S3:** Summary Statistics

|  | CA, NY,<br>WA | 95% conf.<br>interval |  | Other<br>States | 95% conf.<br>interval |  | Used Fox<br>News | 95% conf.<br>interval |  | Used<br>CNN | 95% conf.<br>interval |  |
| --- | --- | --- | --- | --- | --- | --- | --- | --- | --- | --- | --- | --- |
| 1) Get Covid | **.208** | .191 | .225 | **.200** | .189 | .210 | **.174** | .159 | .189 | **.228** | .212 | .244 |
| 2) Die from Covid | **.133** | .119 | .147 | **.140** | .130 | .150 | **.146** | .132 | .160 | **.143** | .129 | .157 |
| 3) Excess Mortality | **.036** | .030 | .042 | **.039** | .035 | .043 | **.036** | .030 | .041 | **.045** | .039 | .052 |
| 4) Lose Job | **.124** | .106 | .142 | **.093** | .081 | .104 | **.099** | .084 | .114 | **.105** | .090 | .119 |
| 5) Out of Money | **.164** | .146 | .182 | **.123** | .112 | .134 | **.139** | .123 | .155 | **.144** | .127 | .161 |
| 6) Distancing | **.776** | .747 | .805 | **.672** | .651 | .693 | **.684** | .653 | .715 | **.783** | .756 | .809 |

*Notes:* The table shows the mean and 95% confidence intervals by state levels of infection and by source of information of 1) the chances of getting the virus within three months, 2) the chances of dying from the virus if infected, 3) excess mortality, 4) the chances of losing job within three months, 5) the chances of running out of money because of the virus within three months and 6) whether individuals refrain from at least one social activity. High-infected states include California (CA), New York (NY) and Washington (WA). We use sample weights to make the survey representative of the U.S. population aged 18 and older. Data come from “Understanding America Study” (UAS) collected between March 10 and March 16, 2020.

**Figure S1:**
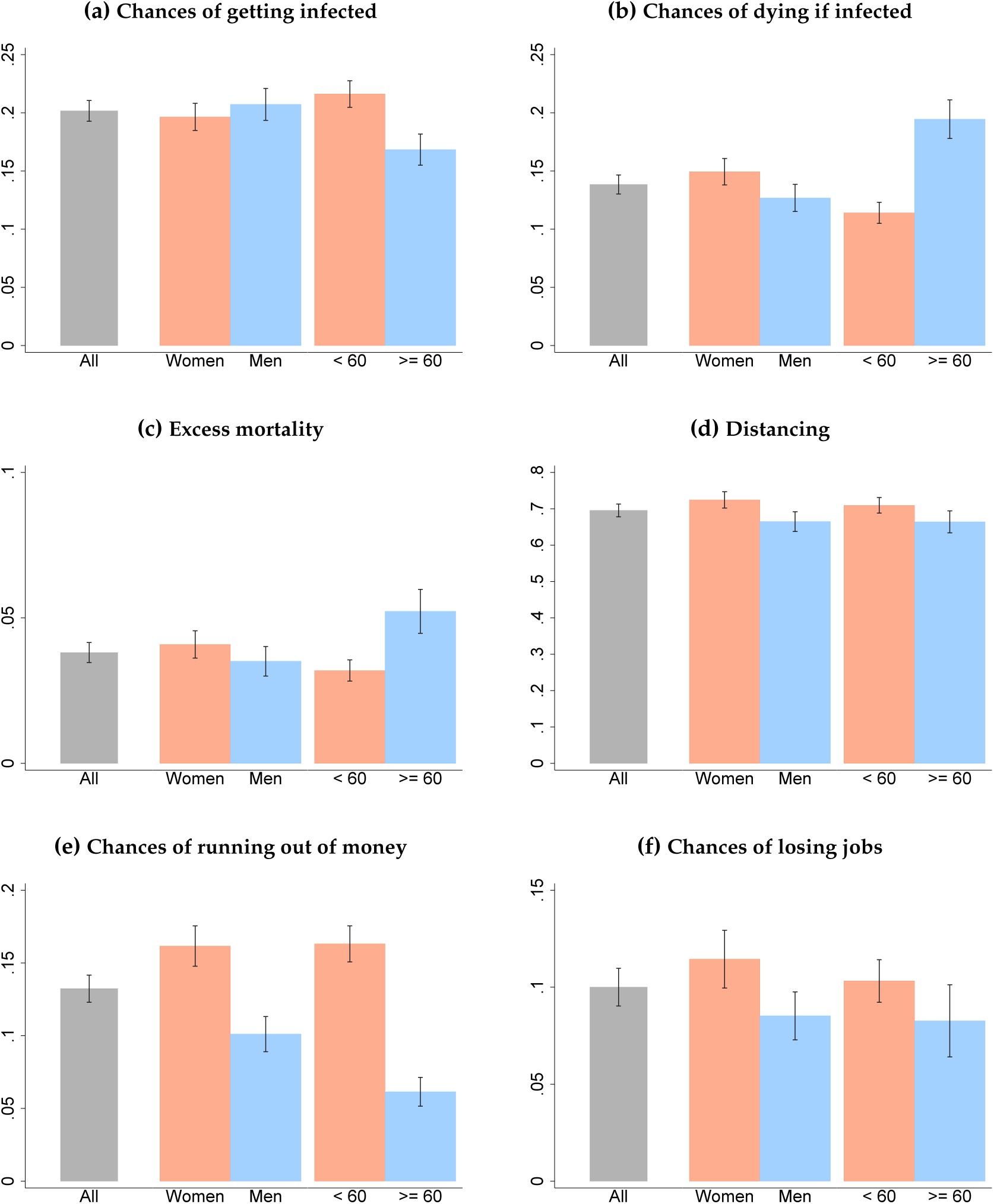
Differences by age and gender. *Notes:* Data coming from “Understanding America Study” (UAS) collected between March 10 and March 16, 2020. Bar plots show weighted means for the six variables we consider in our analysis along with their confidence intervals, derived based on weighted standard errors. We present these statistics for all U.S. residents, and by gender and age categories (below or above 60).

**Table S4:** Heterogeneity

|  | (1)<br>Get Covid | (2)<br>Die from Covid | (3)<br>Excess Mortality | (4)<br>Lose Job | (5)<br>Out of Money | (6)<br>Distancing |
| --- | --- | --- | --- | --- | --- | --- |
| Man | 0.015<br>(0.009) | -0.030***<br>(0.008) | -0.007**<br>(0.004) | -0.022**<br>(0.010) | -0.041***<br>(0.009) | -0.063***<br>(0.018) |
| Age | -0.002***<br>(0.000) | 0.002***<br>(0.000) | 0.001***<br>(0.000) | -0.001***<br>(0.000) | -0.003***<br>(0.000) | -0.001<br>(0.001) |
| High school | 0.010<br>(0.021) | 0.007<br>(0.023) | 0.002<br>(0.011) | -0.006<br>(0.034) | -0.051*<br>(0.027) | -0.035<br>(0.047) |
| Some college | 0.032<br>(0.020) | -0.032<br>(0.021) | -0.007<br>(0.010) | -0.031<br>(0.032) | -0.079***<br>(0.025) | -0.055<br>(0.044) |
| College+ | 0.077***<br>(0.020) | -0.076***<br>(0.020) | -0.020**<br>(0.010) | -0.057*<br>(0.031) | -0.132***<br>(0.024) | 0.018<br>(0.044) |
| Constant | 0.237***<br>(0.022) | 0.075***<br>(0.023) | 0.026***<br>(0.009) | 0.210***<br>(0.038) | 0.369***<br>(0.028) | 0.778***<br>(0.050) |
| Observations | 5274 | 5272 | 5271 | 3248 | 5303 | 5289 |

*Notes:* The table reports the coefficients of linear regressions to show heterogeneity by demographic groups in 1) the chances of getting the virus within three months, 2) the chances of dying from the virus if infected, 3) excess mortality, 4) the chances of losing job within three months, 5) the chances of running out of money because of the virus within three months and 6) whether individuals refrain from at least one social activity. The demographic variables are age, gender and four education categories. We use sample weights to make the survey representative of the U.S. population aged 18 and older. Robust standard errors are reported in parentheses. \* $p < 0.1$ , \*\* $p < 0.05$ , \*\*\* $p < 0.01$ . Data on perceptions and social distancing come from “Understanding America Study” (UAS) collected between March 10 and March 16, 2020.

**Table S5:** Heterogeneity by Source of Information

|  | (1)<br>Get Covid | (2)<br>Die from Covid | (3)<br>Excess Mortality | (4)<br>Lose Job | (5)<br>Out of Money | (6)<br>Distancing |
| --- | --- | --- | --- | --- | --- | --- |
| used Fox News | -0.037***<br>(0.009) | -0.009<br>(0.009) | -0.012***<br>(0.004) | -0.003<br>(0.010) | 0.008<br>(0.014) | -0.023<br>(0.019) |
| used CNN | 0.041***<br>(0.010) | 0.014**<br>(0.007) | 0.014***<br>(0.003) | 0.005<br>(0.008) | 0.015*<br>(0.008) | 0.120***<br>(0.018) |
| Observations | 5223 | 5222 | 5221 | 3214 | 5255 | 5247 |

*Notes:* The table reports the coefficients of linear regressions to show heterogeneity by source of information in 1) the chances of getting the virus within three months, 2) the chances of dying from the virus if infected, 3) excess mortality, 4) the chances of losing job within three months, 5) the chances of running out of money because of the virus within three months and 6) whether individuals refrain from at least one social activity. Used Fox News and CNN are 0/1 answers to the question "Which of the following information sources have you used to learn about the coronavirus in the past 7 days?". Control variables include age, gender, four education categories, state and day fixed effects. We use sample weights to make the survey representative of the U.S. population aged 18 and older. Robust standard errors are reported in parentheses. \* $p < 0.1$ , \*\* $p < 0.05$ , \*\*\* $p < 0.01$ . Data on perceptions and social distancing come from "Understanding America Study" (UAS) collected between March 10 and March 16, 2020.

**Table S6:**
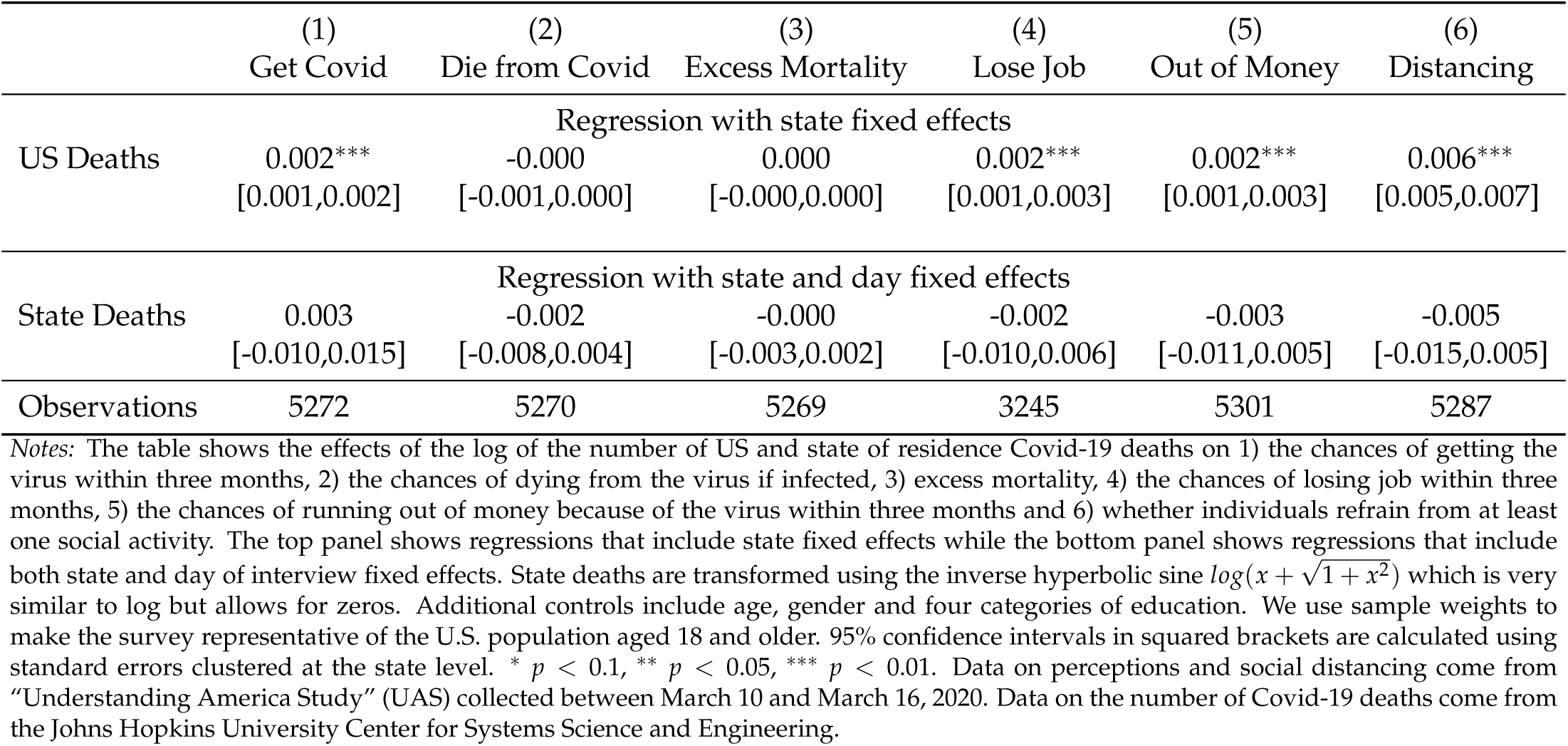
Effects of Covid cumulative deaths on Covid perceptions and social distancing

## Notes

### Competing Interest Statement

The authors have declared no competing interest.

### Funding Statement

We gratefully acknowledge the support of the Population Aging Research Center (PARC) and Population Studies Center (PSC) at the University of Pennsylvania, which are funded by the U.S. National Institutes of Health through grants NIA P30 AG12836 and NICHD R24 HD044964 respectively.

